# Economics of Implementing Preemptive School Closures to Mitigate Pandemic Influenza Outbreaks of Differing Severity in the United States

**DOI:** 10.1101/2021.11.23.21266745

**Authors:** Lori R Dauelsberg, Brian Maskery, Heesoo Joo, Timothy C Germann, Sara Y Del Valle, Amra Uzicanin

**Affiliations:** Analytics, Intelligence and Technology Division, Los Alamos National Laboratory, PO Box 1663, Los Alamos, NM 87545 United States; Division of Global Migration and Quarantine, Centers for Disease Control and Prevention, 1600 Clifton Road NE, MS H16-4, Atlanta, GA 30329 United States; Theoretical Division, Los Alamos National Laboratory

**Keywords:** Community mitigation, economics, influenza, nonpharmaceutical interventions, pandemic, preemptive school closures, societal costs

## Abstract

The use of nonpharmaceutical interventions (NPIs) to slow disease spread, is a part of national pandemic preparedness as the first line of defense against influenza pandemics. Preemptive school closures (PSCs), an NPI reserved for use in severe pandemics, are highly effective in slowing influenza spread but have unintended consequences. We simulated PSC impacts during a 1957-like pandemic to estimate population impacts and quantify costs of closing schools at the national level. We also simulated 1957-like, 1968-like, and 2009-like pandemics at the Chicago regional level. We estimated economic impacts resulting from loss of income due to illness, providing childcare during closures, and other PSC costs while taking into consideration the number of cases averted with each mitigation strategy. The estimated net PSC costs ranged from $15 billion to $192 billion (2016 USD) (1957-like, national-level) where between 2.3 and 47 million US cases may be averted depending on strategy. We found that 2-week school-by-school PSCs (as opposed to county-wide or school district–wide ones) had the lowest cost per discounted life-year gained for both national and Chicago regional-level analyses of all pandemics. While feasibility of such spatiotemporally precise triggering is presently questionable for most locales, this is, theoretically, an attractive option early in an outbreak, while assessing transmissibility and severity of a novel influenza virus. In contrast, we found that county-wide PSCs of longer durations (8 to 12 weeks) would result in the most averted cases (31-47 million) and deaths (105,000-156,000), albeit at considerably more cost ($125-$150 billion net of averted illness costs) for the national-level, 1957-like analysis. The estimated net costs per death averted ($1.0 to $1.2 million) for these scenarios compare favorably to the range of values recommended for regulatory impact analyses ($4.6 to 15.0 million). Hence, economic benefits of such PSCs would exceed the population impacts and economic costs.

## 1. Introduction

Influenza pandemics occur after a novel, readily transmissible influenza virus emerges and spreads rapidly across the globe (Webster et al., 2013). They are inherently unpredictable with regard to timing—over the past 100 years, only 4 have occurred—and vary greatly with regard to transmissibility and clinical severity (Webster et al., 2013).^1^ Community mitigation is part of the national pandemic response plan as the first line of defense for influenza pandemics until pandemic influenza vaccines are widely available (Department of Health and Human Services, 2017). Community mitigation refers to nonpharmaceutical interventions (NPIs), which are a set of actions that people and communities can take to slow the spread of disease (Qualls et al., 2017). Preemptive school closures (PSCs), a community NPI reserved for use in pandemics, are implemented before disease becomes widespread in schools and communities and may be recommended during severe, very severe, and extreme influenza pandemics to achieve one or more of the following specific public health objectives (Qualls et al., 2017):

- **1:** To gain time for an initial assessment of transmissibility and clinical severity of the pandemic virus in the very early stage of its circulation in humans (closures up to 2 weeks).
- **2:** To slow down the spread of the pandemic virus in areas that are beginning to experience local outbreaks and thereby allow time for the local health care system to prepare additional resources for responding to increased demand for health care services (closures up to 6 weeks).
- **3:** To allow time for pandemic vaccine production and distribution (closures up to 6 months – the duration is related with the presently anticipated influenza vaccine production and distribution timelines).

This recommendation is based on a 2012 statement by the U.S. Community Preventive Services Task Force (CPSTF) that recommended coordinated PSCs and dismissals during a severe influenza pandemic (a pandemic with high rates of severe illness such as that experienced in 1918) based on sufficient evidence of effectiveness in reducing or delaying the spread of infection and illness within communities (CPSTF, 2012). However, the CPSTF found insufficient evidence to determine the balance of benefits and harms of preemptive, coordinated school dismissals in the event of an influenza pandemic of moderate or less severity because few studies provide comparative information relevant to an overall assessment of potential benefits and costs of school dismissals for pandemics without high rates of severe illness (CPSTF, 2012) (Community Preventive Services Task Force (CPSTF), 2012) (Community Preventive Services Task Force (CPSTF), 2012) (Community Preventive Services Task Force (CPSTF), 2012) (Community Preventive Services Task Force (CPSTF), 2012).

In this study, we present an economic evaluation from the societal perspective of PSCs implemented during a hypothetical influenza pandemic similar in magnitude to the influenza pandemic in 1957 at the national level as well as 1968-like and 2009-like for the Chicago region (Germann et al., 2019). While the 1957 influenza pandemic was substantially less severe than that of 1918, a contemporary pandemic severity analysis shows it as the most impactful modern-time influenza pandemic (Reed et al., 2013). Given this constellation of factors, we chose to present the 1957-like pandemic severity scenario for the national-level economic evaluation here and include additional scenarios evaluated in the Supplemental Information. Our analysis explores the national economic impacts for the scenario of greatest severity, for which coordinated PSCs are most likely to be relevant both at the state and at the national level. We also explore a larger set of assumptions and pandemic scenarios based on a set of regional “deep dive” epidemiologic model simulations (included in the supplemental information). The goal of this study was to explore costs associated with PSCs as a countermeasure aimed to slow down or lessen the pandemic-associated morbidity and mortality, compared with the cost of a pandemic without this mitigation measure.

## 2. Methods

To estimate the economic impacts of PSCs based on the 1957-like pandemic, we used the pertinent parameter assumptions and model-produced epidemiologic data from a previously reported national-level 1957-like pandemic simulation. The parameters carried over from that work into the present analysis are summarized in Table 1 and Supplemental Information Tables A1-A2 The results of the epidemiologic model included the total number of pandemic influenza cases averted, if the schools are closed, as well as the number of schools closed and the number of students affected (i.e., out of school) (Germann et al., 2019). We used these results as input parameters in the present analysis to determine the total costs of illness, including hospitalizations, for averted cases and the costs to close schools.

**Table 1.**
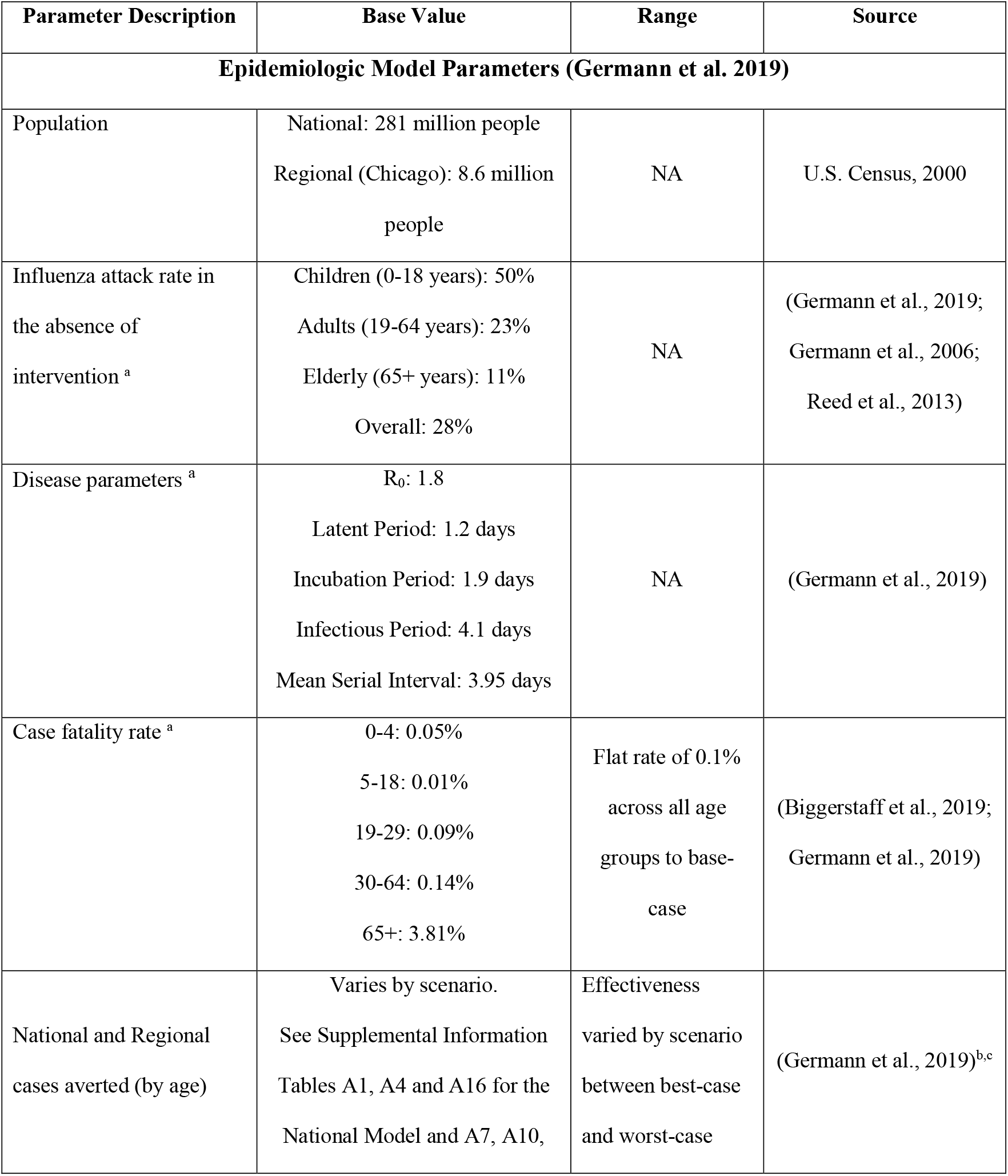
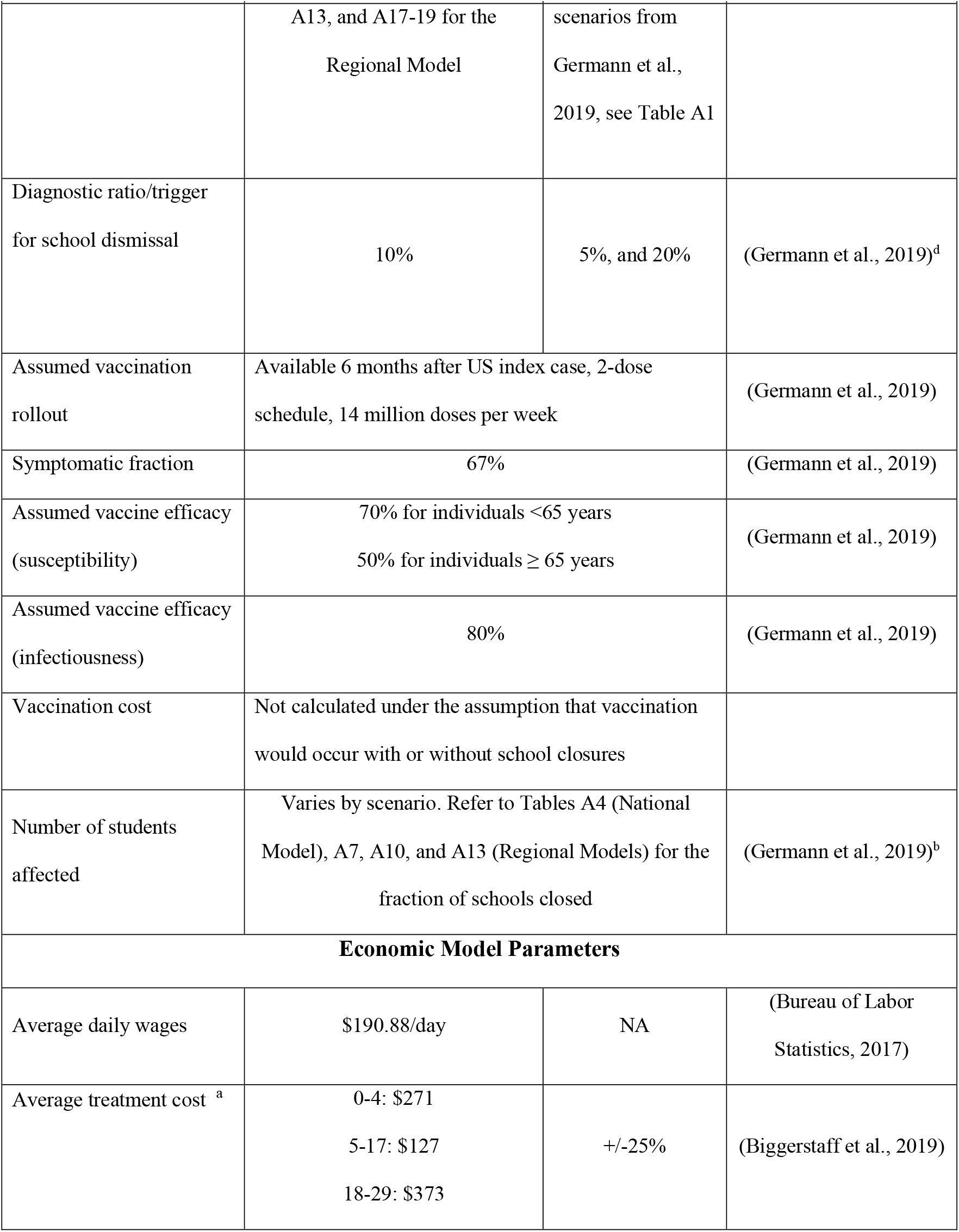
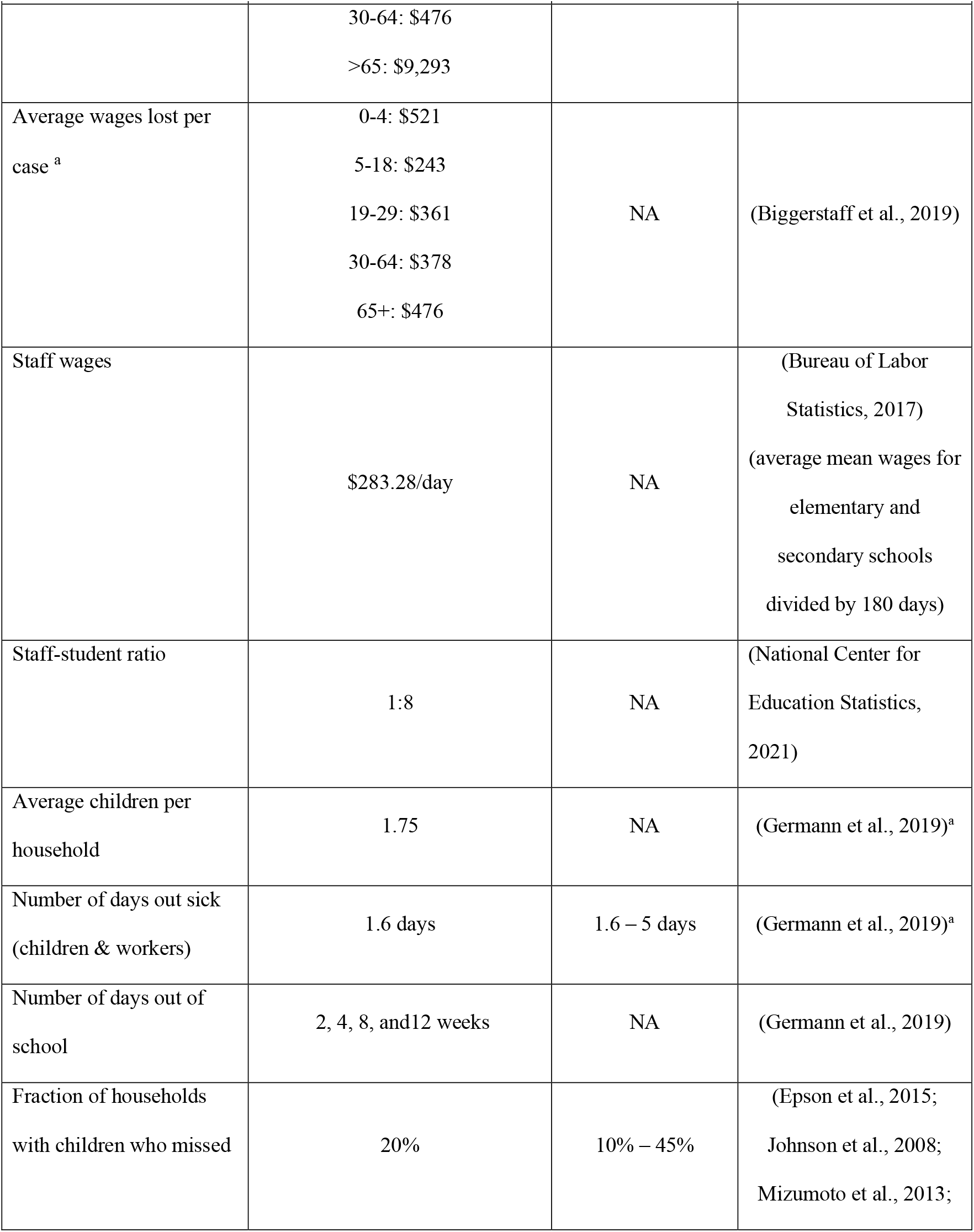
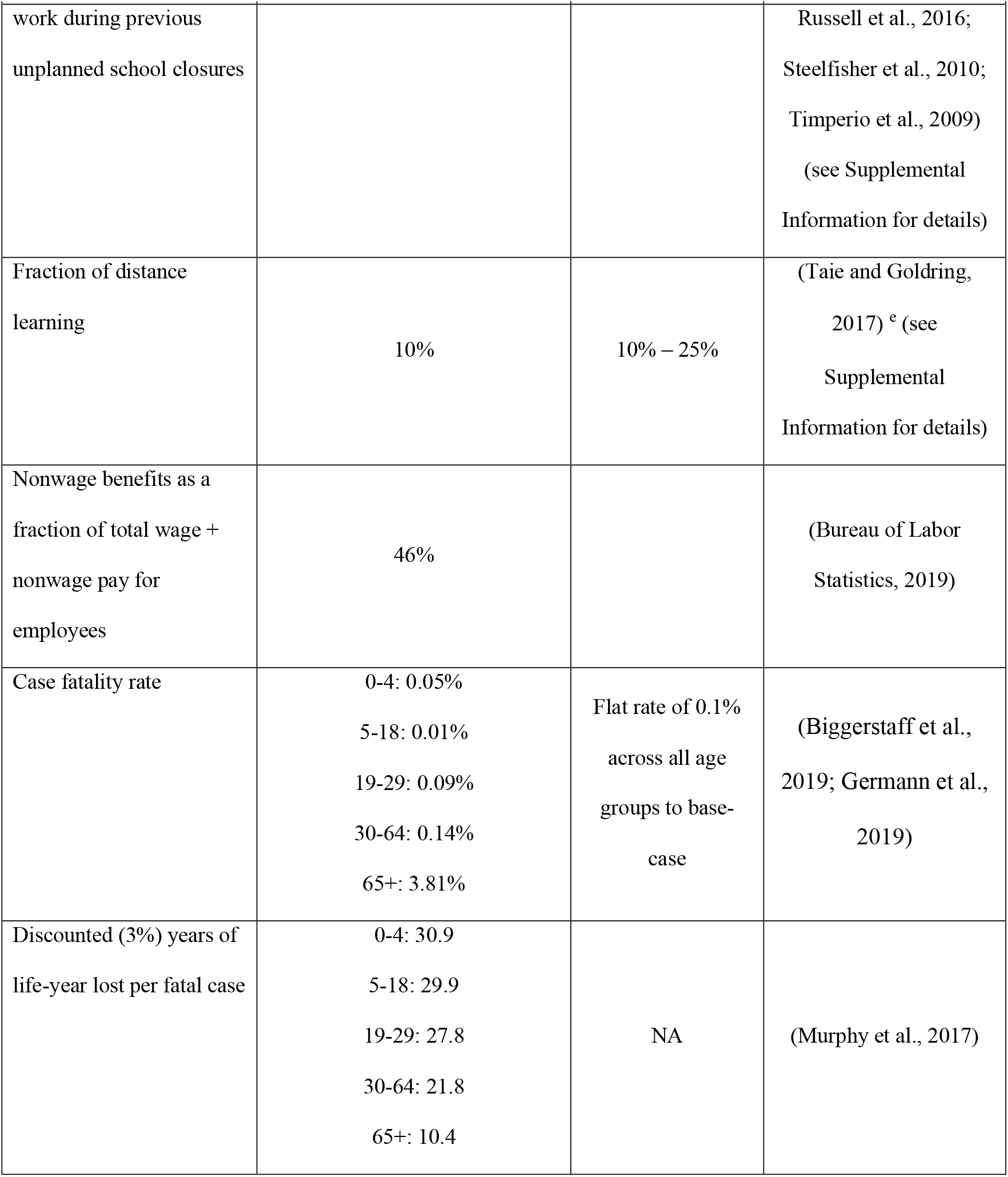
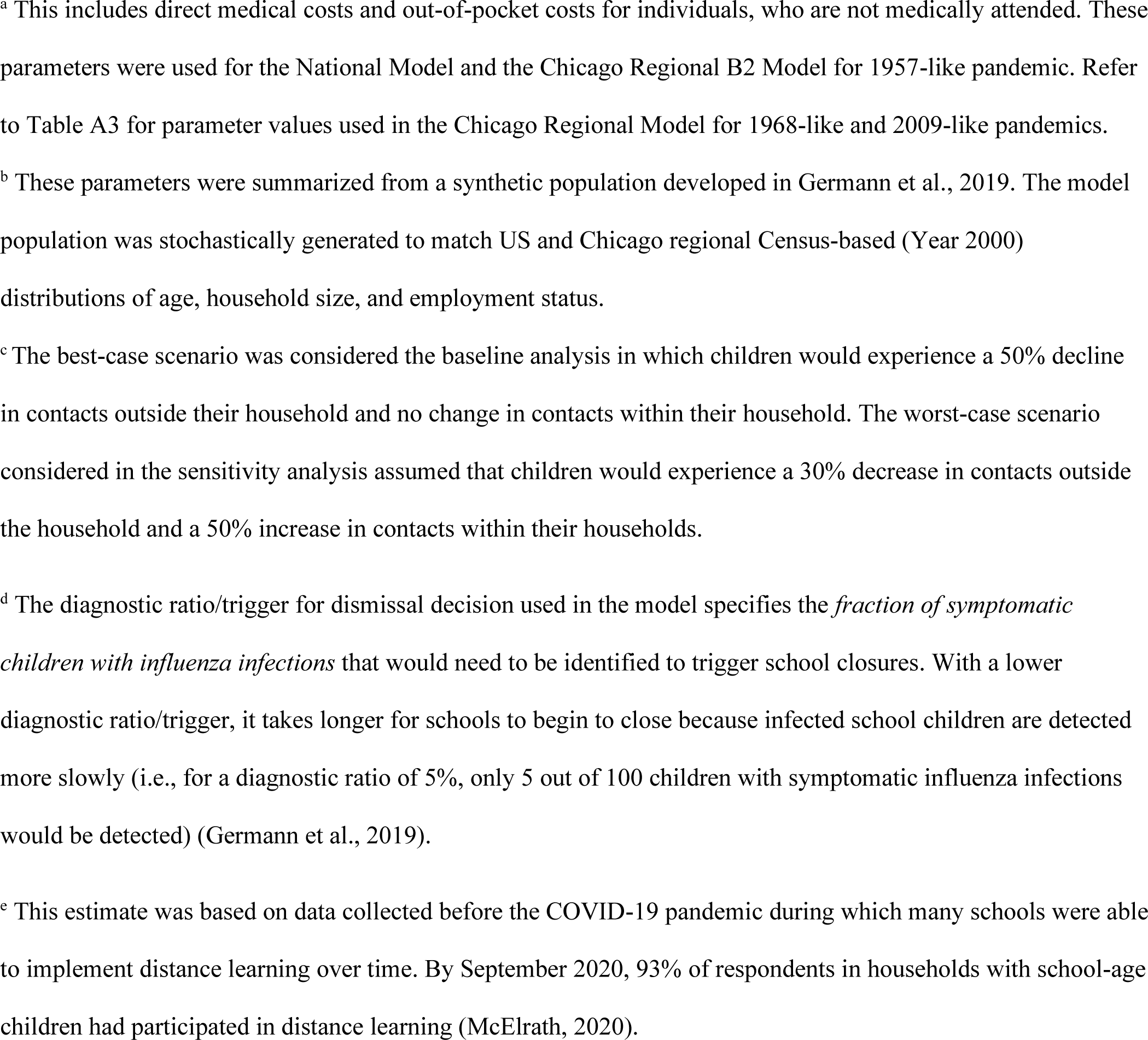
Parameter Values and Assumptions Informing an Economic Analysis of Implementing Preemptive School Closures to Mitigate Pandemic Influenza Outbreaks].

| Parameter Description | Base Value | Range | Source |
| --- | --- | --- | --- |
| **Epidemiologic Model Parameters (Germann et al. 2019)** |  |  |  |
| Population | National: 281 million people<br>Regional (Chicago): 8.6 million people | NA | U.S. Census, 2000 |
| Influenza attack rate in the absence of intervention <sup>a</sup> | Children (0-18 years): 50%<br>Adults (19-64 years): 23%<br>Elderly (65+ years): 11%<br>Overall: 28% | NA | (Germann et al., 2019;<br>Germann et al., 2006;<br>Reed et al., 2013) |
| Disease parameters <sup>a</sup> | R<sub>0</sub>: 1.8<br>Latent Period: 1.2 days<br>Incubation Period: 1.9 days<br>Infectious Period: 4.1 days<br>Mean Serial Interval: 3.95 days | NA | (Germann et al., 2019) |
| Case fatality rate <sup>a</sup> | 0-4: 0.05%<br>5-18: 0.01%<br>19-29: 0.09%<br>30-64: 0.14%<br>65+: 3.81% | Flat rate of 0.1%<br>across all age groups to base-case | (Biggerstaff et al., 2019;<br>Germann et al., 2019) |
| National and Regional cases averted (by age) | Varies by scenario.<br>See Supplemental Information Tables A1, A4 and A16 for the National Model and A7, A10, | Effectiveness varied by scenario between best-case and worst-case | (Germann et al., 2019)<sup>b,c</sup> |
|  | A13, and A17-19 for the<br>Regional Model | scenarios from<br>Germann et al.,<br>2019, see Table A1 |  |
| Diagnostic ratio/trigger<br>for school dismissal | 10% | 5%, and 20% | (Germann et al., 2019)<sup>d</sup> |
| Assumed vaccination<br>rollout | Available 6 months after US index case, 2-dose<br>schedule, 14 million doses per week |  | (Germann et al., 2019) |
| Symptomatic fraction | 67% |  | (Germann et al., 2019) |
| Assumed vaccine efficacy<br>(susceptibility) | 70% for individuals <65 years<br>50% for individuals ≥ 65 years |  | (Germann et al., 2019) |
| Assumed vaccine efficacy<br>(infectiousness) | 80% |  | (Germann et al., 2019) |
| Vaccination cost | Not calculated under the assumption that vaccination<br>would occur with or without school closures |  |  |
| Number of students<br>affected | Varies by scenario. Refer to Tables A4 (National<br>Model), A7, A10, and A13 (Regional Models) for the<br>fraction of schools closed |  | (Germann et al., 2019)<sup>b</sup> |
| Economic Model Parameters |  |  |  |
| Average daily wages | \$190.88/day | NA | (Bureau of Labor<br>Statistics, 2017) |
| Average treatment cost <sup>a</sup> | 0-4: \$271<br>5-17: \$127<br>18-29: \$373 | +/-25% | (Biggerstaff et al., 2019) |
| | 30-64: \$476<br>>65: \$9,293 | | |
| Average wages lost per case <sup>a</sup> | 0-4: \$521<br>5-18: \$243<br>19-29: \$361<br>30-64: \$378<br>65+: \$476 | NA | (Biggerstaff et al., 2019) |
| Staff wages | \$283.28/day | NA | (Bureau of Labor Statistics, 2017)<br>(average mean wages for elementary and secondary schools divided by 180 days) |
| Staff-student ratio | 1:8 | NA | (National Center for Education Statistics, 2021) |
| Average children per household | 1.75 | NA | (Germann et al., 2019)<sup>a</sup> |
| Number of days out sick (children & workers) | 1.6 days | 1.6 – 5 days | (Germann et al., 2019)<sup>a</sup> |
| Number of days out of school | 2, 4, 8, and 12 weeks | NA | (Germann et al., 2019) |
| Fraction of households with children who missed | 20% | 10% – 45% | (Epson et al., 2015; Johnson et al., 2008; Mizumoto et al., 2013; |
| work during previous<br>unplanned school closures |  |  | Russell et al., 2016;<br>Steelfisher et al., 2010;<br>Timperio et al., 2009)<br>(see Supplemental<br>Information for details) |
| Fraction of distance<br>learning | 10% | 10% – 25% | (Taie and Goldring,<br>2017) <sup>e</sup> (see<br>Supplemental<br>Information for details) |
| Nonwage benefits as a<br>fraction of total wage +<br>nonwage pay for<br>employees | 46% |  | (Bureau of Labor<br>Statistics, 2019) |
| Case fatality rate | 0-4: 0.05%<br>5-18: 0.01%<br>19-29: 0.09%<br>30-64: 0.14%<br>65+: 3.81% | Flat rate of 0.1%<br>across all age<br>groups to base-case | (Biggerstaff et al.,<br>2019; Germann et al.,<br>2019) |
| Discounted (3%) years of<br>life-year lost per fatal case | 0-4: 30.9<br>5-18: 29.9<br>19-29: 27.8<br>30-64: 21.8<br>65+: 10.4 | NA | (Murphy et al., 2017) |

<sup>a</sup> This includes direct medical costs and out-of-pocket costs for individuals, who are not medically attended. These parameters were used for the National Model and the Chicago Regional B2 Model for 1957-like pandemic. Refer to Table A3 for parameter values used in the Chicago Regional Model for 1968-like and 2009-like pandemics.
<sup>b</sup> These parameters were summarized from a synthetic population developed in Germann et al., 2019. The model population was stochastically generated to match US and Chicago regional Census-based (Year 2000) distributions of age, household size, and employment status.
<sup>c</sup> The best-case scenario was considered the baseline analysis in which children would experience a 50% decline in contacts outside their household and no change in contacts within their household. The worst-case scenario considered in the sensitivity analysis assumed that children would experience a 30% decrease in contacts outside the household and a 50% increase in contacts within their households.
<sup>d</sup> The diagnostic ratio/trigger for dismissal decision used in the model specifies the *fraction of symptomatic children with influenza infections* that would need to be identified to trigger school closures. With a lower diagnostic ratio/trigger, it takes longer for schools to begin to close because infected school children are detected more slowly (i.e., for a diagnostic ratio of 5%, only 5 out of 100 children with symptomatic influenza infections would be detected) (Germann et al., 2019).
<sup>e</sup> This estimate was based on data collected before the COVID-19 pandemic during which many schools were able to implement distance learning over time. By September 2020, 93% of respondents in households with school-age children had participated in distance learning (McElrath, 2020).

Briefly, these epidemiological inputs are from an agent-based computational model, also known as Epidemiological Forecasting (EpiCast), designed to simulate community-level influenza transmission in the United States at the national scale (Germann et al., 2019). The national-scale simulation model consists of 281 million individuals distributed among 65,334 census tracts to closely represent the actual population distribution according to publicly available 2000 US Census data (Germann et al., 2019). Each tract is in turn organized in 2,000-person communities resulting in 180,492 model communities. The model combines US Census demographics and worker-flow data to generate daytime and evening contact networks based on potential contacts emerging at schools, workplaces, households, neighborhoods, and communities (Germann et al., 2019).

The national model uses 3 geographic scales for PSC decisions including “community,” “county,” and “multi-county,” which correspond to closing schools one at a time (i.e., community), for the entire school district (i.e., county), or for a group of adjoining counties (i.e., multi-county) once the dismissal decision is reached for any given school. This means that the multi-county closure area is the most aggressive decision to prevent the spread of the pandemic in terms of the number of schools closed. Closure duration scenarios included 2 weeks, 4 weeks, 8 weeks, and 12 weeks. Our study considered the cost of each of these intervention strategies as simulated by the EpiCast model (Germann et al., 2019). The model also examined different triggers for school dismissal based on the fraction of symptomatic children that would cause closures to occur. The trigger is linked to assumptions about the sensitivity of the surveillance system used to detect infected schoolchildren based on the diagnostic ratio. The diagnostic ratio is based on the fraction of symptomatic children infected with influenza who would be detected. With a lower diagnostic ratio/trigger, it takes longer for schools to begin to close because infected school children are detected more slowly (i.e., for a diagnostic ratio of 5%, only 5 out of 100 children with symptomatic influenza virus infections would be detected). This paper focused on an assumed diagnostic ratio of 10% and provided estimates for 5% and 20% diagnostic ratios in the sensitivity analyses and supplemental information.

### 2.1 Net Economic Cost of Preemptive School Closures

We compute the (net) economic cost of the intervention based on the total cost of PSCs minus the averted cost of pandemic cases due to PSCs. The cost of PSCs was derived from the wage losses of school staff due to the schools closing and the wage losses of parents who must care for children out of school. The wage losses of school staff from schools closing include costs for school employees, who are assumed not to work during the closure. The economic costs include lost productivity even if the teachers are paid during the closure. We developed a proxy to estimate this lost productivity based on the wages of school employees multiplied by the duration of school closure. We applied a correction factor (base-case: 10%, range: 0% to 20%) called *fraction of distance learning* to account for schools that are capable of providing education through online learning to offset some of the lost productivity caused by the school closure based on the pre-COVID-19 baseline (see supplemental information).

To estimate the cost of parents’ lost wages, we first divided the number of affected children by the average number of children per household (1.75) to estimate the number of affected households. The number of households with children was then multiplied by the *fraction of households with children whose parents missed work* to estimate the number of parents missing work. Next, the estimated number of parents missing work was multiplied by the US average hourly wage rate with non-wage benefits, the duration of school closures, and an assumption of 8 hours worked per day.

Not all parents would need to miss work to care for children during PSCs. To estimate the *fraction of households with children whose parents missed work*, we reviewed published studies that reported the fraction of parents that missed work due to unplanned school closures in response to the 2009 H1N1 pandemic or due to local excess absenteeism from elevated numbers of influenza-like-illness in US communities (see supplemental information). These studies found that adults from between 14% and 29% of households had missed time at work due to school closures (Epson et al., 2015; Johnson et al., 2008; Mizumoto et al., 2013; Russell et al., 2016; Steelfisher et al., 2010; Timperio et al., 2009). One of the studies was a national survey of households impacted by school closures during the 2009 H1N1 pandemic and found that adults in 20% of households had at least one parent who missed work (Table A2). Among households in which an adult had to miss work, we assumed that one parent or guardian from each family would be at home to care for the child(ren). The base-case scenario assumed that 20% of households (range 10% to 40%) would have at least one parent who would miss time at work to care for children during PSCs. We used a conservative estimate by assuming that one adult in each of these households would miss work during the full period of the school PSC. The sensitivity analysis as well as the regional “deep dive” explores variations of this parameter (see supplemental information). This factor helps to account for families that include older children or other non-working adults (e.g., retired grandparents) that would be able to supervise younger children as well as non-working parents or parents with access to telework, staggered work schedules, or other means to reduce the economic costs to households from lost time at work.

The effectiveness of each intervention was calculated based on the percentage reduction in the expected number of cases relative to no intervention as reported previously (Germann et al., 2019). The averted cost of pandemic cases from PSCs includes treatment costs of cases averted, productivity loss of ill workers associated with averted cases, and productivity loss of parents of ill children. The treatment costs were defined to include direct medical costs incurred from visiting healthcare providers and estimated out-of-pocket costs for non-medically attended cases. The age-specific treatment costs of cases averted were estimated based on a recent analysis of the costs of pandemic influenza in the United States using insurance claims data (Biggerstaff et al., 2019). The productivity loss of ill workers was estimated for numbers of cases averted among the working adult and senior populations. The cases averted for the working adult and senior populations were multiplied by average daily wages (the US average hourly wage rate with an assumption of 8 hours worked per day considering non-wage benefits) and number of days out because of illness. The productivity loss of parents of ill children was estimated by multiplying numbers of cases averted due to PSC for children 12 years old or younger, fraction of working adult population, average daily wages including non-wage benefits, and number of days out due to taking care of ill children. All costs are presented in 2016 US dollars (USD). More details are provided in the supplemental information.

We estimated the numbers of deaths averted based on the numbers of cases averted as a benefit of the intervention. The number of deaths averted was calculated by multiplying the number of cases averted by the age-specific case fatality rate (Biggerstaff et al., 2019). Using these age-specific case fatality rates results in a higher overall case fatality rate than the 0.1% rate used in Germann et al. (2019). This assumption was made because the present age distribution in the United States is considerably older than in 1957, when this pandemic occurred, and the case fatality rate in the >65 years age group was much higher than for other age groups (3.81%). The number of life-years gained is calculated by multiplying the number of deaths averted by age group by the average U.S. life expectancy for each age group (Murphy et al., 2017) after discounting future life-years using a 3% annual rate.

The cost-effectiveness of the intervention was assessed calculating the net cost per case/death averted and the net cost per life-years gained. The net economic cost of the intervention was divided by the change in health outcomes to calculate each ratio. The cost effectiveness ratios were calculated relative to no intervention and incrementally. Incremental cost effectiveness ratios were calculated to examine the marginal cost and the marginal change in health outcomes in moving from shorter durations of closures to longer duration for a given strategy. The relative efficiency of interventions is evaluated based on whether the cost per change in health outcome averted is higher or lower for one intervention strategy compared to another such that the intervention with a lower cost per change in health outcome is considered more efficient (or more cost-effective).

We also analyzed the impact of PSCs for three hypothetical influenza pandemics with viruses with transmissibility and clinical severity similar to those of the 1957 (referred to as 1957-like), 1968 (1968-like), and 2009 (2009-like) influenza pandemics using a regional model of the Chicago region as previously reported (Germann et al., 2019). The input parameters for these models are generally similar to those of the national-level model, except for accounting for different attack rates and severity for each pandemic influenza virus and using a regional rather than national model of the population impacted. Key parameters are summarized in Tables 1 and A3. The regional analysis only considered 2 geographic scales: individual schools (i.e., community) and regional (i.e., multi-county). The regional model is similar to the multi-county approach used in the national analysis.

However, since the Chicago region itself is one multi-county region, in practice, the regional approach would lead to the simultaneous closure of all schools once triggered.

### 2.2 Sensitivity Analysis

We conducted multiple univariate sensitivity analyses to account for uncertainties in the cost of PSCs, the value of averted cases, the effectiveness of PSCs in reducing or delaying the spread of infection, and the case fatality rate. The estimated effect of each parameter on the net cost of the intervention was plotted to demonstrate the relative importance of uncertainty resulting from each parameter. The effectiveness of PSCs varied from a “worst-case” impact on transmission with a doubling of assumed contacts among individuals within a household and only a 30% reduction in contacts among children across households relative to a “best-case” scenario. In the best-case scenario, there is no increase in household contacts and a 50% reduction in contacts among children from different households is assumed. The “best-case” scenario is shown as the “base-case” scenario in this analysis.

## 3. Results

Focusing first on the PSCs that were triggered under the 10% diagnostic ratio assumption for the national model (1957-like), the net costs of the PSC increased from $15 billion for 2-week community-level closures to $192 billion for 12-week multi-county-level closures (Figure 1a). The net costs increased significantly with the duration of the closure and as the geographic scale of closures increases. The benefits in terms of the number of cases and deaths averted through PSCs varied from 2.3 million cases and 7,100 deaths averted for the 2-week multi-county closures to 47 million cases and 156,000 deaths averted for the 12-week county-level closures (Table A4). The duration of closure had a larger impact on the number of cases averted for both the county and multi-county closures areas than for the community closures. For example, in doubling the closure duration from 4 weeks to 8 weeks, the number of cases averted increased by 2.6 and 4.2 times for the county and multi-county closures, but only by 1.6 times for the community closures.

**Figure 1a.**
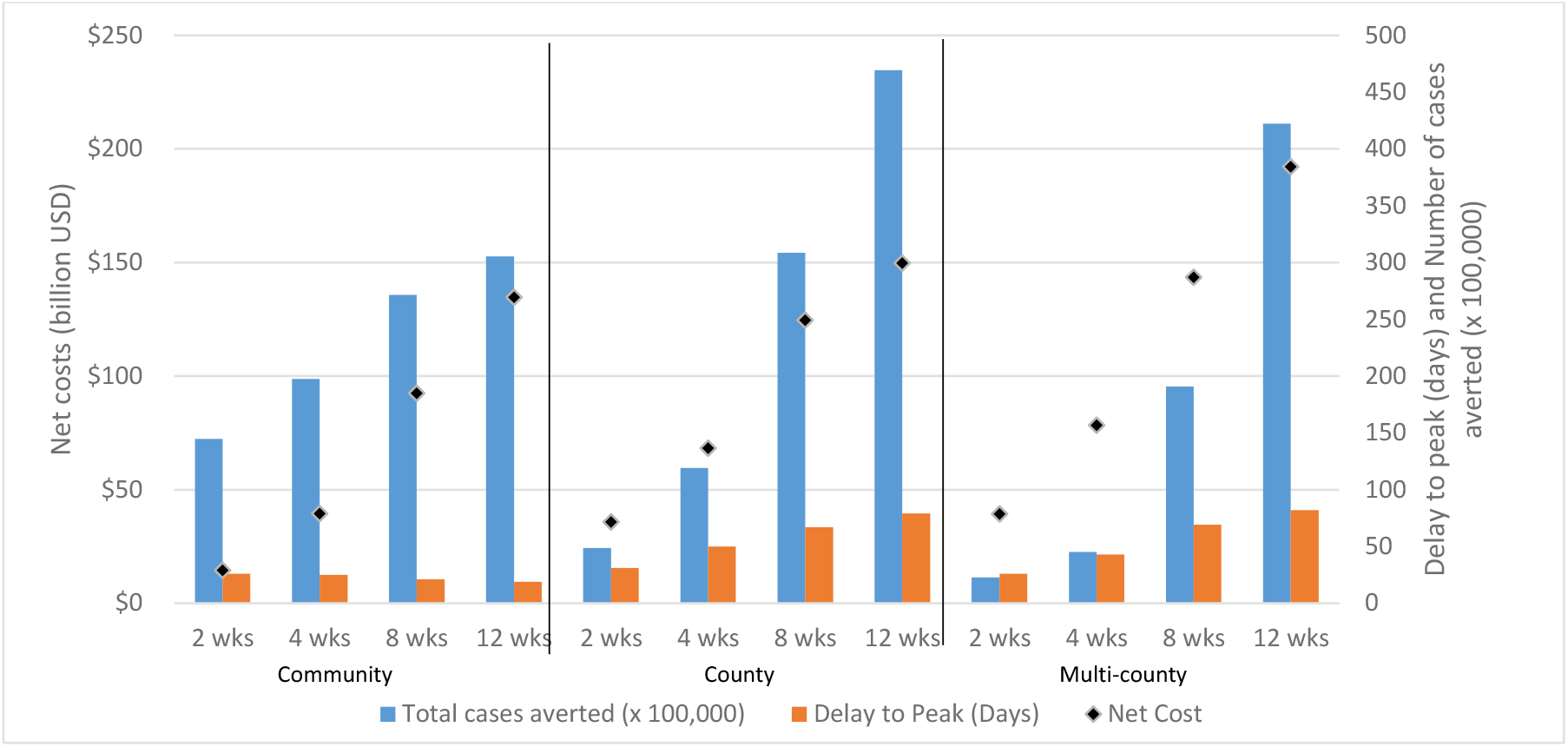
Effect of increasing the **duration of school closures** (while keeping constant the geographic scale of school closures) on cases averted, delay to peak, and net cost for a national-level strategy against an influenza pandemic similar to the 1957 pandemic^*^ ^*^This figure is based on the 10% diagnostic ratio for school closures under the assumption that only 10% of symptomatic school children infected with influenza would be detected by a symptom-based surveillance system and schools would close when the first symptomatic child was diagnosed (Germann et al., 2019).

Figure 1b shows that the net cost was consistently lower for the community-level closures for an equivalent duration and that the number of cases averted was highest for the community closures for durations less than or equal to 4 weeks, but highest for county-level closures for durations of eight weeks or greater. The multi-county-level closures consistently had the highest costs and fewest cases averted for a given duration with the exception of the 12-week duration for which the number of cases averted was greater than for the community closures. For durations greater than 2 weeks, the community closures had less impact on the delay to peak incidence relative to county or multi-county closures. The longest delay to peak (82 days) resulted from a 12-week, multi-county closure strategy. This compares to 79 days for a 12-week county closure strategy or 19 days for a 12-week community closure strategy.

**Figure 1b.**
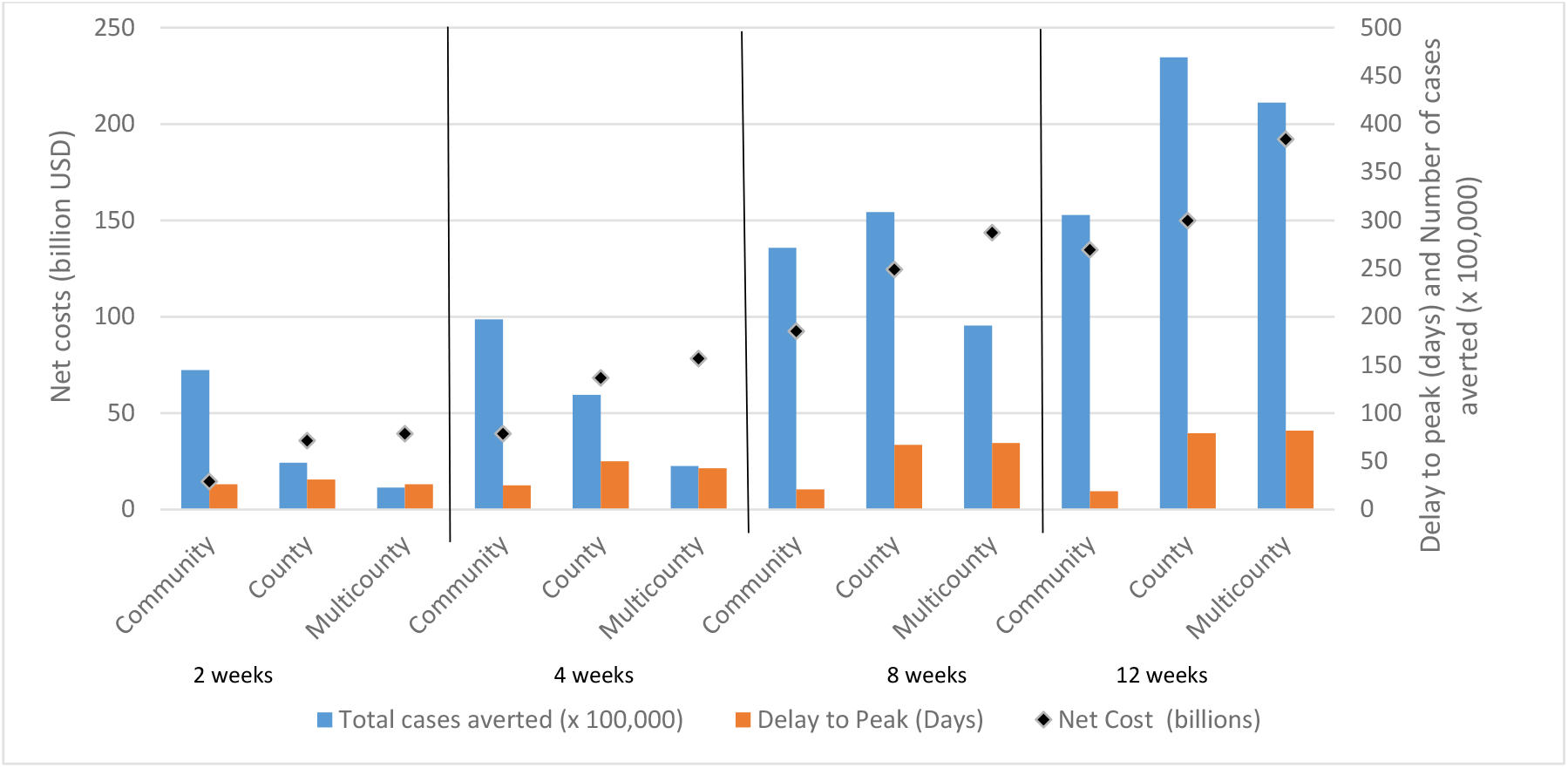
Effect of increasing the **geographic scale** of school closures (while keeping constant the duration) on cases averted, delay to peak, and net cost for a national-level strategy against an influenza pandemic similar to the 1957 pandemic^*^ ^*^This figure is based on the 10% diagnostic ratio for school closures under the assumption that only 10% of symptomatic school children infected with influenza would be detected by a symptom-based surveillance system and schools would close when the first symptomatic child was diagnosed (Germann et al., 2019). Note Multicounty = Multi-county

The net costs and numbers of life-years gained for each intervention on the national scale are shown in Figure 2 and the net cost per discounted life-year gained for each intervention are summarized in Supplemental Information Tables A4-A6. The most efficient intervention was the 2-week community-level closure, with a net cost per discounted life-year gained of about $27,000 relative to no intervention. This would be the best alternative to support an initial assessment of the transmissibility and clinical severity of the pandemic virus (Objective 1 above). For community closures, the net cost per discounted life-year gained increased with duration from $27,000 for 2 weeks to $123,000 for 12-week closures (Supplemental Information Table A5). The incremental cost per discounted life year gained from increasing the duration of community closures increased from $130,000 (4 weeks vs. 2 weeks) to $388,000 (12 weeks vs. 8 weeks) (Supplemental Information Table A6). This demonstrates decreasing returns to scale for increasing the duration of community closures. In contrast, the net cost per discounted life-year gained decreased from 2-week closures ($175,000 for county-level and $435,000 for multi-county–level) to 12-week closures ($76,000 for county and $108,000 for multi-county) such that 12-week durations for both county and multi-county closures would be more efficient than community closures (Supplemental Information Tables A5 and A6). The increasing returns to scale suggest that county closures became more efficient in terms of the net cost per discounted life-year gained with increasing duration from 2 weeks to 12 weeks. The number of cases averted and life years gained were maximized with 12-week county-level closures (Objective 3 above).

**Figure 2.**
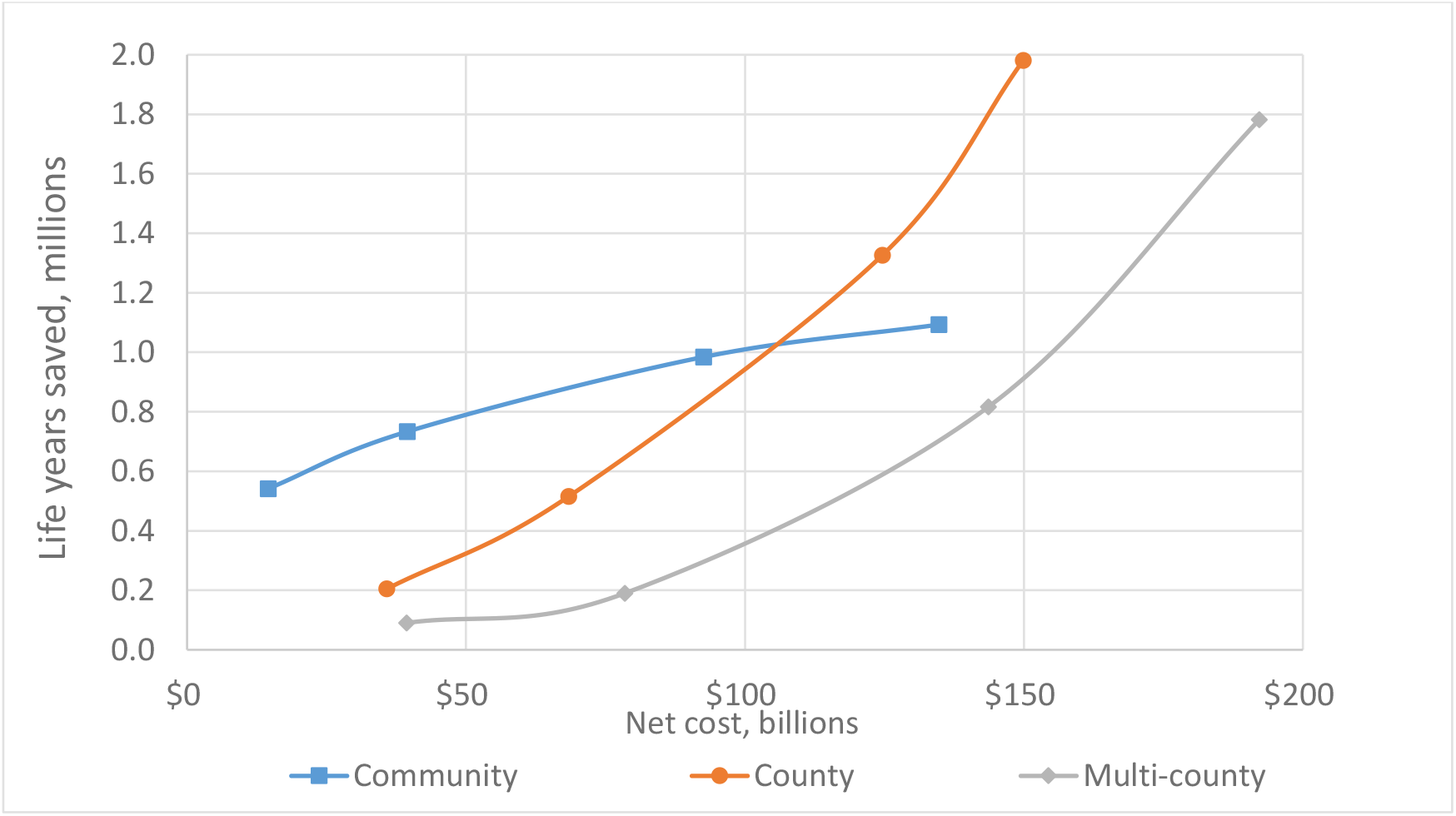
**Net cost** (billion USD) and **number of life-years gained** for community-, county-, and multi-county– level school closures implemented nationwide during a hypothetical influenza pandemic similar to the 1957 influenza A (H2N2) pandemic^*^ Notes: The duration of school closures increases from 2 weeks to 4 weeks to 8 weeks to 12 weeks for community-level closures (blue), county-level closures (orange), and multi-county-level closures (gray). The net costs and life-years gained increase with the durations of closures for each geographic scale. ^*^This figure is based on the 10% diagnostic ratio for school closures under the assumption that only 10% of symptomatic school children infected with influenza would be detected by a symptom-based surveillance system and schools would close when the first symptomatic child was diagnosed (Germann et al., 2019).

A series of one-way sensitivity analyses of the net cost per case averted, per death averted, and per life-year gained are shown in Figures 3a through 3d. Relative to county-level closures with a 4-week duration, shorter durations were less costly but also less efficient, and longer durations were more costly and more efficient. At 4 weeks, a shift from county-to community-level closures would increase efficiency, while a shift to multi-county– level PSCs would reduce efficiency (i.e., increase the net cost per case/death averted or life-year gained). The decision trigger/diagnostic ratio had less effect on effectiveness or efficiency relative to duration or geographic scale. The assumed diagnostic ratio had the greatest impact on community closures (Supplemental Information Table A4 and Figures A1a-c and A2a-c) such that the 20% trigger was much more effective and the 5% trigger much less effective than the 10% trigger.

**Figure 3a.**
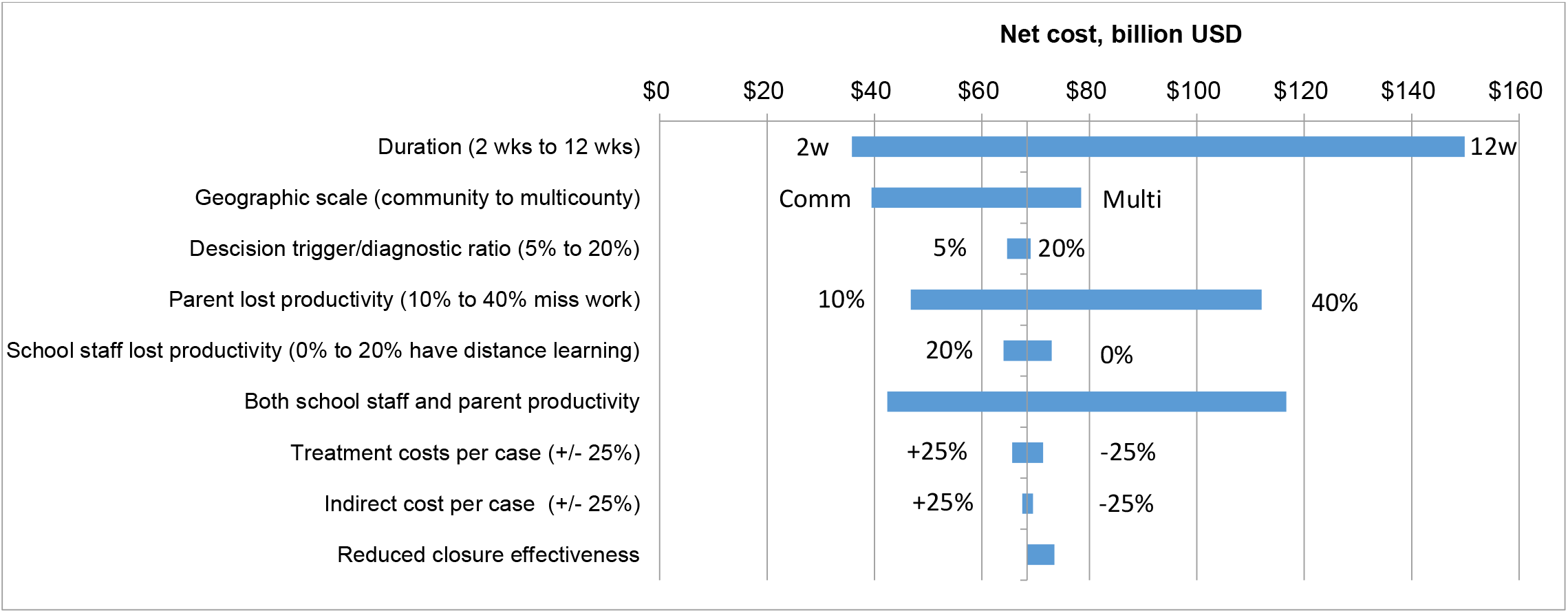
One-way sensitivity analyses of the **net cost of school closure** to the 4-week county-level closure with a 10% diagnosis ratio assumption^*^ at base-case values Baseline estimates: Duration 4 weeks; Geographic scale County; Parent lost productivity 20%; School staff lost productivity 20%; $69 billion net cost ^*^This figure is based on the 10% diagnostic ratio for school closures under the assumption that only 10% of symptomatic school children infected with influenza would be detected by a symptom-based surveillance system and schools would close when the first symptomatic child was diagnosed (Germann et al., 2019). Treatment costs were defined to include direct medical costs incurred from visiting healthcare providers and estimated out-of-pocket costs for non-medically attended cases

**Figure 3b.**
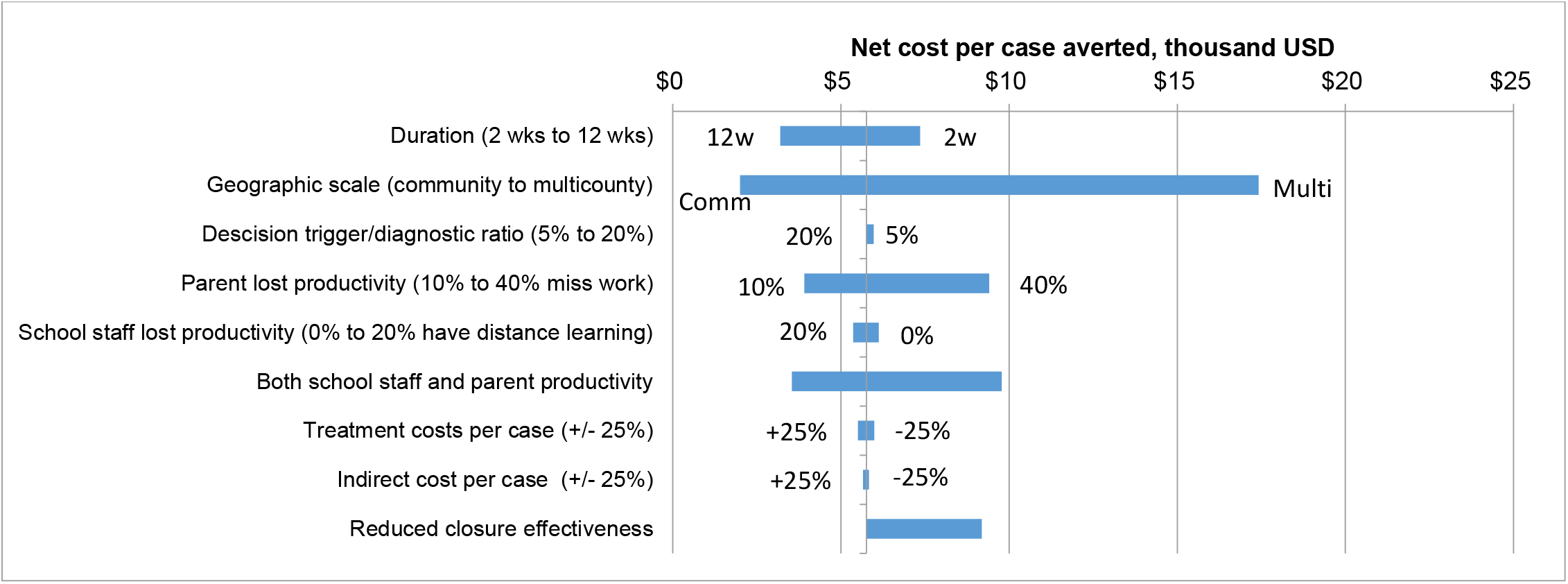
One-way sensitivity analyses of the **net cost per case averted** relative to the 4-week county-level closure with a 10% diagnostic ratio assumption^*^ at base-case values Baseline estimates: Duration 4 weeks; Geographic scale County; Parent lost productivity 20%; School staff lost productivity: 20%; $5,760 per case averted ^*^This figure is based on the 10% diagnostic ratio for school closures under the assumption that only 10% of symptomatic school children infected with influenza would be detected by a symptom-based surveillance system and schools would close when the first symptomatic child was diagnosed (Germann et al., 2019). Treatment costs were defined to include direct medical costs incurred from visiting healthcare providers and estimated out-of-pocket costs for non-medically attended cases.

**Figure 3c.**
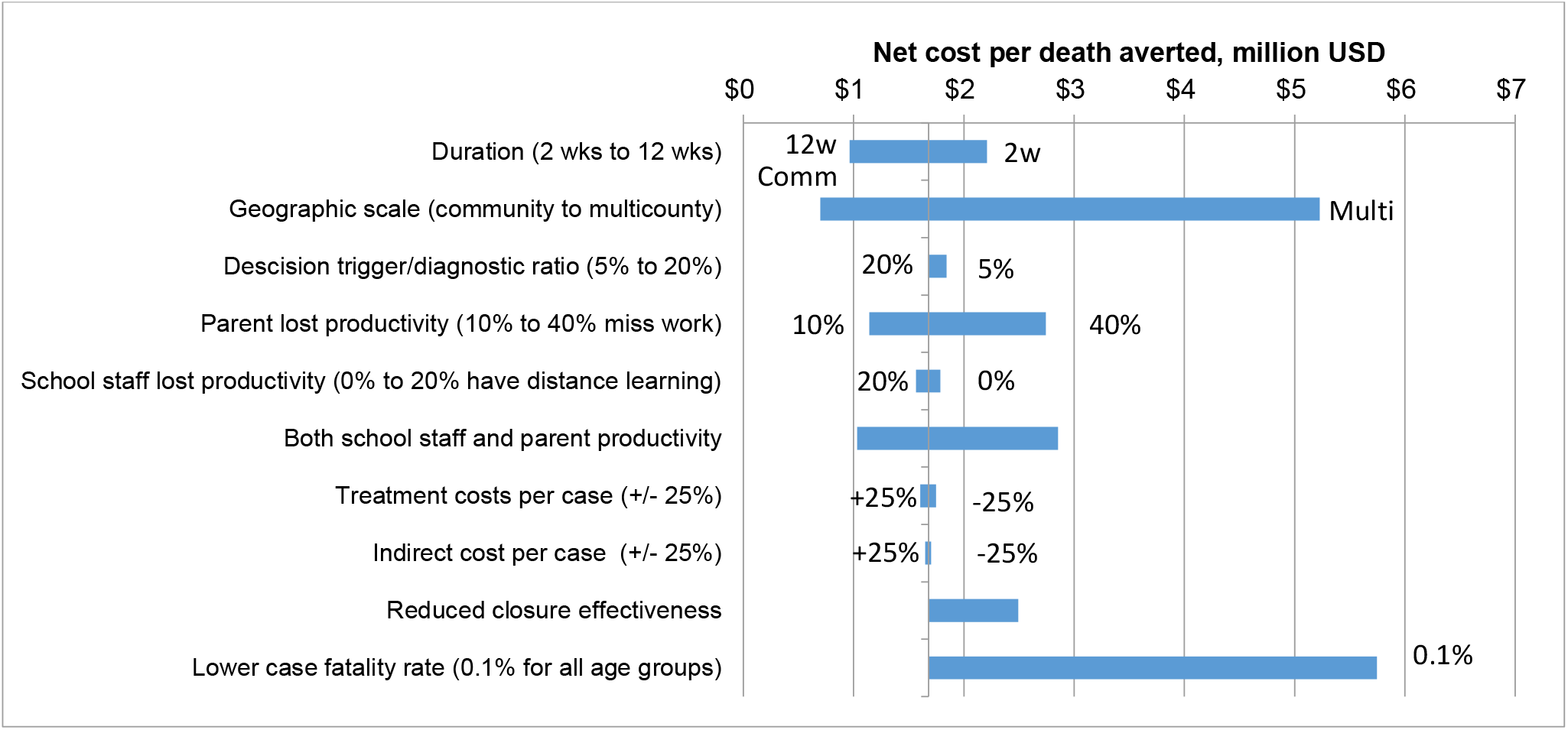
One-way sensitivity analyses of the **net cost per death averted** relative to the 4-week county-level closure with a 10% diagnosis ratio assumption^*^at base-case values Baseline estimates: Duration 4 weeks; Geographic scale County; Parent lost productivity 20%; School staff lost productivity 20%; $1.68 million per death averted ^*^This figure is based on the 10% diagnostic ratio for school closures under the assumption that only 10% of symptomatic school children infected with influenza would be detected by a symptom-based surveillance system and schools would close when the first symptomatic child was diagnosed (Germann et al., 2019). Treatment costs were defined to include direct medical costs incurred from visiting healthcare providers and estimated out-of-pocket costs for non-medically attended cases.

**Figure 3d.**
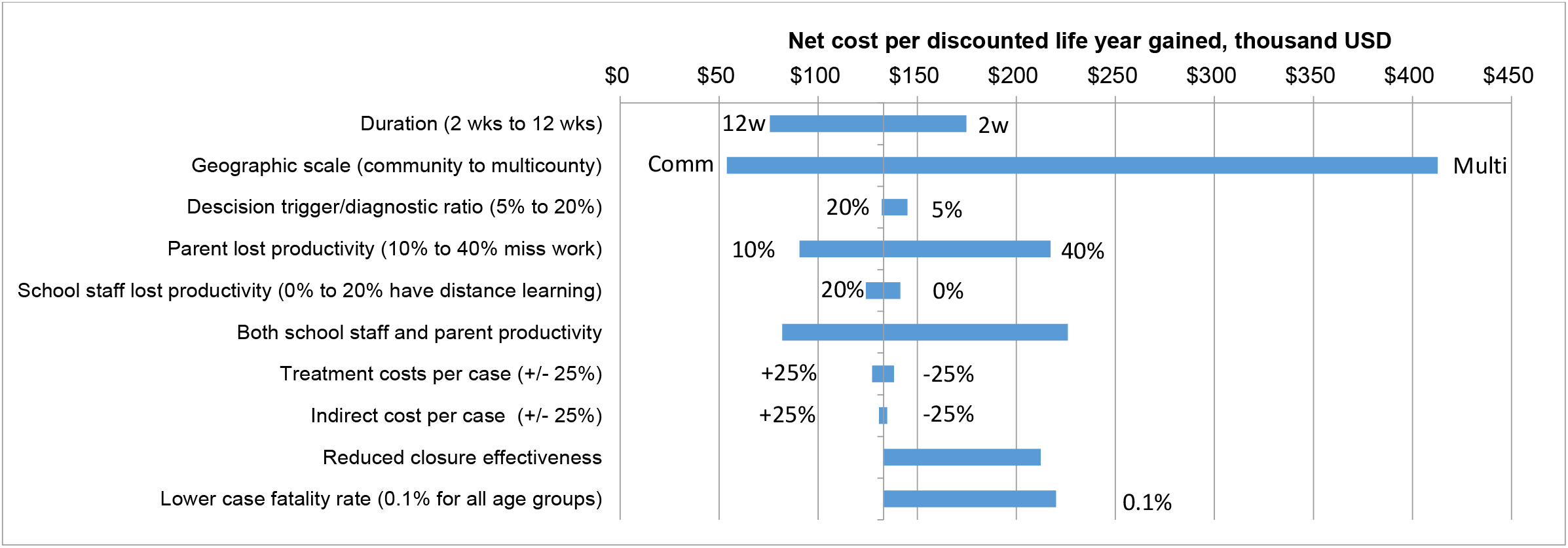
One-way sensitivity analyses of the **net cost per life-years gained** relative to the 4-week county-level closure with a 10% diagnosis ratio assumption^*^ at base-case values Baseline estimates: Duration 4 weeks; Geographic scale County; Parent lost productivity 20%; School staff lost productivity 20%; $133,000 per life-year gained ^*^This figure is based on the 10% diagnostic ratio for school closures under the assumption that only 10% of symptomatic school children infected with influenza would be detected by the surveillance system and schools would close when the first symptomatic child is diagnosed (Germann et al., 2019). Treatment costs were defined to include direct medical costs incurred from visiting healthcare providers and estimated out-of-pocket costs for non-medically attended cases.

Lost productivity for parents during PSCs resulted in more uncertainty than the fraction of schools with distance learning capabilities, which affected lost productivity of school staff. The net cost for a 4-week, county-level closure varied from $47 billion to $112 billion (base-case: $68 billion) depending on the fraction of parents that would miss time at work due to the closures. In contrast, uncertainty in the fraction of schools with distance learning capacity only resulted in a range of $64 to $73 billion. This was due in part to the limited capacity for distance learning (only 10% of schools at the pre-COVID 19 baseline). In comparison to the costs of closure, the uncertainty in averted costs (benefits) from averted illnesses contributed to less uncertainty in net cost and efficiency estimates.

If PSCs were less effective than assumed in the base-case, the net cost per case averted would increase from $5,800 to $9,200, the net cost per death averted would increase from $1.7 million to $2.5 million, and net cost per discounted life-year gained would increase from $133,000 to $212,000 for a 4-week, county-level closure. Another source of uncertainty in the net cost per death averted and per discounted life-year gained was the case fatality rate such that the net cost per death averted would increase to $5.8 million and the net cost per discounted life-year gained would increase to $220,000 if a constant 0.1% case fatality rate were applied to all age groups.

For the Chicago regional analysis, the findings for the 1957-like influenza pandemic were similar to those for the national-level model except that the estimated costs per life-year gained were considerably higher (e.g., Chicago region: net cost of $60,000 to $144,000 per discounted life-year gained for community-level closures with 10% dismissal trigger versus $27,000 - $124,000 for the national model). The net cost per discounted life-year gained from multi-county-level (regional) closures with 10% dismissal trigger in the Chicago area exceeded $1.3 million for all durations less than 12 weeks. For the 1968-like and 2009-like pandemics, the number of life-years gained and savings from averted cases were lower than for the 1957-like pandemic because of lower transmissibility and severity for these pandemics. The community closures resulted in lower net costs than the regional closures for all three pandemics and the net cost per discounted life-year gained for community-level closures with 10% dismissal trigger exceeded $100,000 for all interventions except the 2-week community closures for the 2009-like pandemic. Refer to Supplemental Information, Tables A7-A15 and Figures A1a-c, A3a-b, and A5a-b, A6, and A7.

## 4. Discussion

There are important considerations to the decisions made in closing schools preemptively in response to an influenza pandemic. With larger geographic scale and longer durations of PSCs, their costs dramatically increase. The net costs of closures with 10% dismissal trigger vary from $15 billion for 2-week community-level (school-by-school) closures to $192 billion for 12-week multi-county-level closures. However, as the costs of closures increase, the benefits in terms of cases and deaths averted also increase from 2.3 million cases and 7,100 deaths averted for 2-week multi-county closures to 47 million cases and 156,000 deaths averted for 12-week county closures. This congruence of the directions between increasing costs and in parallel increasing benefits of preemptive school closures underscores the importance of optimizing the duration and geographic scale of preemptive closures for the desired public health objective. The net cost per death averted varied between $350,000 and $5.6 million. In comparison, the U.S. Department of Health and Human Services (HHS) has recommended that estimates of between $4.6 and $15.0 million (2016 USD) of the value of statistical life be used in regulatory impact analyses (Department of Health and Human Services, 2016). The net cost per death averted for all strategies, with the exception of multi-county closures ≤4 weeks, was less than the recommended lower-bound value of statistical life, indicating that mortality reduction benefits of PSCs would exceed the estimated costs of closures for a pandemic similar to that which occurred in 1957.

In our model, county-level closures approximate school district-level closures, because most US counties have a single public K-12 school district. Compared to community-level (i.e., single school) closures, the county closures are equally or more cost-efficient (in terms of net cost per death averted) for durations ≥8 weeks and maximize the number of cases and deaths averted (Figure 2). Additionally, in the United States, school district-wide closures are much more frequent than closures of individual schools (Wong et al., 2014; Zviedrite et al., 2019), likely reflecting the programmatic considerations as well as the organizational level that usually authorizes such a drastic measure. In general, both county- and multi-county-level closures are associated with greater delays to peak incidence (i.e., flattening the curve), albeit at higher net costs than community closures. County closures have lower net costs as well as more cases/deaths averted compared to multi-county closures with similar though slightly shorter delays to peak incidence.

Community-level closures were estimated to be considerably more cost-efficient in terms of the net cost per case or death averted for shorter durations (≤4 weeks). This geographic scale of closure in our model is most congruent to school-by-school decision making, where each individual school (rather than an entire school district) would decide on its own to close. In a potential pandemic, this lowest level of PSCs may be both pertinent and appropriate very early in an evolving pandemic; for example, if a school-associated case or cluster of cases of unsubtypable influenza A is recognized quickly, this will prompt a local investigation to determine the epidemiologic features of the outbreak. However, in practice, such a scenario is highly unlikely, since it would require each individual school to have the capability to promptly and accurately detect and diagnose cases of novel (i.e., unsubtypable) influenza A among students and staff. Sensitivity analyses suggest that the number of cases averted would increase with more sensitive surveillance, reflected in a higher trigger (20%) for dismissal decisions for community-level interventions. The net cost per life-year gained would decrease from a range of $27,000 to $123,000 for community closures with a 10% trigger to $13,000 to $41,000 with a 20% trigger. This suggests that sensitive and timely surveillance needed for prompt triggering of the intervention is especially important for the effectiveness and efficiency of the community PSC approach, but less important for the county and multi-county approaches.

The net cost per case averted varied from about $1,000 to $7,500 for most scenarios with the exception of the multi-county–level closures for ≤4 weeks for which the net cost per case averted was greater than $17,000. The net cost per death averted varied from $0.35 to $1.6 million for community-level closures, $0.97 to $2.2 million for county-level closures, and $1.4 to $5.6 million for multi-county-level closures. All of the estimated net costs per death averted fell within or below the range of $4.5 to $14.6 million, which is a recommended estimate of the value of statistical life from the HHS (Department of Health and Human Services, 2016). The net cost per life-year gained varied from $27,000 to $123,000 for community closures, $76,000 to $175,000 for county closures, and $108,000 to $435,000 for multi-county closures. These estimates can be compared to regulatory guidance for the valuation of a quality adjusted life-year gained, which vary from $230,000 to $750,000 (Department of Health and Human Services, 2016).

Not surprisingly, the net cost per life-year gained was much higher for the less severe pandemics (1968-like and 2009-like scenarios) compared to the 1957-like pandemic, as described in the Supplemental Information. Given the significant costs associated with PSCs, while influenza pandemic severity is still unknown, it is prudent to close schools for shorter periods (e.g., 2 weeks) to help gather the data needed to ascertain more information about the specific virus. To the extent possible, during influenza pandemics PSCs should be executed at the smallest possible geographic scales (e.g., individual schools or school districts), because the wider geographic scope (regional / multi-county) and short-term closures (e.g., 2 weeks) were estimated to have limited cost effectiveness for reducing caseloads; the net cost per discounted life-year gained with a 10% dismissal trigger was estimated to be $7.5 million for the 1968-like pandemic and $1.0 million for the 2009-like pandemic. In contrast, the estimated net cost per life-year gained for 2-week, community-level (school-by-school) and school district closures were $226,000 for the 1968-like and $68,000 for 2009-like pandemics.

A limited number of studies have attempted to quantify the potential economic costs associated with school closures for a specific region and/or pandemic severity. For example, Sadique et al. (2008) estimated the potential economic cost associated with school closures during mild to severe pandemics (i.e., ranging from 2 to 12 weeks) in the United Kingdom. Their findings show that closures could result in 16% workforce absenteeism and £0.2 to £1.2 billion costs per week. Lempel et al. (2009) estimated the potential economic costs associated with closing all schools in the United States for 2 to 12 weeks as a response to mild to severe pandemics. Their findings show that closing all schools for a 4-week period could result in $10 to $47 billion dollars in costs among households with school-age children and a reduction of 6% to 19% in key health personnel. The estimated costs to households in this analysis fell within the range estimated by Lempel et al., although overall costs were higher in this analysis because we included costs to school staff. Brown et al. (2011) used an agent-based simulation to explore school closures in Pennsylvania ranging between 1 and 8 weeks during the 2009 H1N1 pandemic. Their findings showed that closing schools for 8 weeks would have resulted in median net costs of $21 billion (95% range: $8 to $45.3 billion). They concluded that the cost associated with school closures might have outweighed the cost savings in preventing influenza cases during the 2009 pandemic. These cost estimates included lost wages for households and lost productivity for school employees and were significantly higher than our cost estimates.

Our findings should be considered in context of several potential limitations. First, the costs of PSCs on the schools and school employees as well as the parents of school children were difficult to estimate because of their unprecedented nature. Specifically, large-scale PSCs had not been attempted in the United States to control infectious disease outbreaks until the COVID-19 pandemic in 2020. We estimated these parameters based on observations from short-term school closures during previous influenza outbreaks and attempted to account for this uncertainty by conducting a sensitivity analysis of costs for schools and the parents of school children. One strength of this analysis is the incorporation of empirical data on the impacts of PSCs on parents’ abilities to continue working even if the data are limited to short-term closures. However, the fraction of households in which parents would miss work to care for children during closures and the fraction of schools with distance learning capabilities were estimated using a pre-COVID-19 baseline. About 93% of people in households with school-age children had participated in distance learning by September, 2020 (Mcelrath, 2020). If these distance learning capabilities are maintained into the future, the economic costs of school closures may be reduced from the estimates presented above. For example, if 75% of schools maintain distance learning capabilities instead of the 10% estimated in the base-case above, the total economic cost of school closures would decrease by 35%. Improvement to telework capabilities could also reduce the costs of parents missing work to care for children during closures.

Second, treatment costs are based on data from more recent influenza outbreaks and estimated differences in hospitalization rates between the 1957 pandemic and more recent outbreaks, which were much less severe than the 1957 pandemic (Biggerstaff et al., 2019). The potential case fatality rate also remains uncertain. On the one hand, the effectiveness of treatment for influenza has improved since 1957. Yet, the age distribution for the US population has shifted upward such that a greater fraction of the total population is in older, a higher risk category for severe influenza illness. In the sensitivity analysis, we analyzed a flat 0.1% case fatality rate across age groups as well as the population-weighted case fatality rates presented in Table 1. However, even with such a relatively low assumed pandemic case-fatality ratio, our results are consistent with other published model-based analyses which have examined hypothetical outbreaks with higher case fatality rates (Araz et al., 2012; Xue et al., 2012). The number of life years lost was estimated based on the average life expectancy by age group; however, the risk of death from influenza increases for patients with medical comorbidities (Paules and Subbarao, 2017), who would also be expected to have shorter life expectancies relative to the average for each age group.

The third limitation is related with the fact that the ability to forecast the spread of a transmissible disease and its severity is limited to data from past pandemics and hence may not be accurate. This study relies on historic information and data associated with only one particular virus of pandemic influenza, the 1957 influenza A (H2N2), for the national model, and three pandemic influenza viruses (1957-like, 1968-like, and 2009-like) for the regional model, and a limited sensitivity analysis around the effectiveness of school closures. In the baseline analysis, children were assumed to reduce their contacts outside the household by 50% (30% in the sensitivity analysis) with no change in contacts within the household (household contacts were doubled in the sensitivity analysis). One strength of the present analysis is the use of an agent-based model at the national scale, which incorporates demographic and spatial heterogeneities at sub-regional levels and enables head-to-head comparisons between multiple geographic scales of closures.

Fourth, PSCs also may result in unintended consequences not captured in this study due to the limited data available to parametrize the model. For example, some parents have expressed concerns about arranging childcare during prolonged school closures, which could result in job losses. Academic performance (e.g., standardized test scores) in districts affected by prolonged school closures may be lower, potentially affecting future student placement (Marcotte and Hemelt, 2008). For students enrolled in school meal programs, school closures may cause additional financial burden for their families as some affected students may miss meals that they would get at school, even as the vast majority of parents interviewed (>96%) support the decision to close schools (Timperio et al., 2009). Loss of other school-based services, such as counseling or specialized support for some students with disabilities, may disproportionally impact certain vulnerable groups of students who rely on them. School closures also introduce equity concerns regarding the abilities of different socio-economic groups to cope with closures (Armitage and Nellums, 2020; Lee and Lubienski, 2016), which deserves attention as millions of students in the United States and worldwide had to study online, for prolonged periods of time in 2020-2021, during the COVID-19 pandemic caused by a novel coronavirus, SARS-CoV2. These and any other presumed unintended consequences and potential coping mechanisms that limit their detrimental impact on children and families are beyond the scope of this study due to limited data to inform these potential outcomes.

## 5. Conclusion

We found that closing schools by county (or school district) for longer durations (8 to 12 weeks) would result in the most cases (31-47 million) and deaths (105,000 - 156,000) averted, albeit at considerable cost ($125-$150 billion net of averted illness costs). The net cost per death averted was estimated to be between $1.0 and $1.2 million and the net cost per life-year gained between $76,000 and $94,000 for these scenarios. These estimates compare very favorably to the range of value of statistical life estimates recommended for regulatory impact analyses ($4.6 to 15.0 million) suggesting that the benefits of preemptive school closures to mitigate influenza pandemics of varying severity would exceed the costs. We also found closing schools individually for 2-week periods had the lowest cost per discounted life-year gained ($27,000). The finding supported that community-level closures are an attractive alternative at the outset of an outbreak, while attempting to assess the transmissibility and severity of a new pandemic influenza virus. These estimates can assist decision makers assess the cost and benefits of PSCs in response to influenza pandemics.

## Data Availability

All data produced in the present study are available upon reasonable request to the authors.

## Acknowledgements

The authors wish to thank Nidhi Parikh, Noreen Qualls, and Matthew Biggerstaff for their help with data, reviewing and editing, and support of this publication.

## Footnotes

1 This work is focused on influenza pandemics, and the analysis and writing presented here occurred primarily before the start of the coronavirus diseases 2019 (COVID-19) pandemic in 2020. As such, several references to pandemics and large-scale closing of schools do not take into account what had occurred during that coronavirus-associated pandemic.

